# Co-infection in critically ill patients with COVID-19: An observational cohort study from England

**DOI:** 10.1101/2020.10.27.20219097

**Authors:** Vadsala Baskaran, Hannah Lawrence, Louise Lansbury, Karmel Webb, Shahideh Safavi, Izzah Zainuddin, Tausif Huq, Charlotte Eggleston, Jayne Ellis, Clare Thakker, Bethan Charles, Sara Boyd, Tom Williams, Claire Phillips, Ethan Redmore, Sarah Platt, Eve Hamilton, Andrew Barr, Lucy Venyo, Peter Wilson, Tom Bewick, Priya Daniel, Paul Dark, Adam R Jeans, Jamie McCanny, Jonathan D Edgeworth, Martin J Llewelyn, Matthias L Schmid, Tricia M McKeever, Martin Beed, Wei Shen Lim

## Abstract

**Objective:** To describe the incidence and nature of co-infection in critically ill adults with COVID-19 infection in England.

**Methods:** A retrospective cohort study of adults with COVID-19 admitted to seven intensive care units (ICUs) in England up to 18 May 2020, was performed. Patients with completed ICU stays were included. The proportion and type of organisms were determined at <48 and >48 hours following hospital admission, corresponding to community and hospital-acquired co-infections.

**Results:** Of 254 patients studied (median age 59 years (IQR 49-69); 64.6% male), 139 clinically significant organisms were identified from 83(32.7%) patients. Bacterial co-infections were identified within 48 hours of admission in 14(5.5%) patients; the commonest pathogens were *Staphylococcus aureus* (four patients) and *Streptococcus pneumoniae* (two patients). The proportion of pathogens detected increased with duration of ICU stay, consisting largely of Gram-negative bacteria, particularly *Klebsiella pneumoniae* and *Escherichia coli*. The co-infection rate >48 hours after admission was 27/1000 person-days (95% CI 21.3-34.1). Patients with co-infections were more likely to die in ICU (crude OR 1.78,95% CI 1.03-3.08, p=0.04) compared to those without co-infections.

**Conclusion:** We found limited evidence for community-acquired bacterial co-infection in hospitalised adults with COVID-19, but a high rate of Gram-negative infection acquired during ICU stay.

## Introduction

During previous viral pandemics, reported co-infection rates and implicated pathogens have varied. In the 1918 influenza pandemic an estimated 95% of severe illness and death was complicated by bacterial co-infection, predominantly *Streptococcus pneumoniae* and *Staphylococcus aureus* [1].

As of 3 September 2020, over 25 million cases and 850 000 deaths due to COVID-19 infection have been reported world-wide [2]. The symptoms associated with COVID-19 infection are relatively non-specific. Fever and lower respiratory tract symptoms, such as a cough or breathlessness, are common in patients who require hospital care and radiological changes consistent with pneumonia are evident in up to 97% of these patients [3]. Confirmation of acute COVID-19 infection is reliant on a positive SARS-CoV-2 polymerase chain reaction (PCR) test result. The immune response to SARS-CoV2 infection includes a rise in IL-6 and C-reactive protein (CRP), with higher levels associated with more severe disease [4, 5].

The contribution of secondary or co-pathogens to COVID-19 infection is not well understood. The lack of an effective anti-viral agent against SARS-CoV2 combined with challenges in differentiating secondary bacterial co-infection from severe COVID-19 infection alone, has fostered the widespread use of empirical antibiotics in the immediate management of patients hospitalised with COVID-19 infection. Over the spring wave of the pandemic, 83.1% of hospitalised patients in the UK received empirical antibiotic treatment [6].

The utility of specific biomarkers such as procalcitonin to guide antibiotic therapy in severe respiratory tract infection, and specifically COVID-19 infection, is as yet uncertain [7, 8]. In the meantime, a better understanding of the incidence of co-infection in patients with COVID-19 infection and the pathogens involved is necessary for effective antimicrobial stewardship. The primary objective of this study was to determine the rate of laboratory-proven co-infection in critically ill adults with COVID-19 infection in England. Secondary aims were to describe the organisms, the characteristics of patients with co-infection and the antibiotic susceptibilities of identified bacteria.

## Methods

### Data source

A retrospective observational multicentre study of co-infection in adults with confirmed COVID-19 requiring intensive care unit (ICU) admission was performed. Seven acute hospitals from across England participated in the study including large (>1000 beds) tertiary hospitals and medium (500-1000 beds) district hospitals: Nottingham University Hospitals NHS Trust, Newcastle Upon Tyne Hospitals NHS Foundation Trust, Brighton and Sussex University Hospitals NHS Trust, Guy’s and St Thomas’ NHS Foundation Trust, Salford Royal NHS Foundation Trust, University Hospitals of Derby and Burton NHS Foundation Trust and University College London Hospitals NHS Foundation Trust.

### Study population

Case inclusion criteria were adults aged >16 years with completed ICU admissions (discharged from or died whilst in ICU) for COVID-19 pneumonia (i.e. requiring Level 2 or Level 3 care according to the classification by the Intensive Care Society, UK) from disease emergence to 18 May 2020. SARS-CoV-2 was confirmed using reverse transcriptase-polymerase chain reaction (RT-PCR) from a respiratory specimen. Participating sites were asked to enter data for either: 1) all identified patients, or 2) a random selection of at least ten patients from across their eligible cohort. Where more than one critical care area existed at a participating site, a random selection from across areas was requested to avoid selection bias. Exclusion criteria were defined as: COVID-19 infection diagnosed >48 hours after hospital admission or a hospital admission in the last 14 days (hospital-acquired COVID-19) and patients transferred into ICU from a different hospital. Only the first admission to ICU was included.

### Data collection

Personal information was removed at the point of participating site data entry onto a secure online database platform (REDCap Cloud). Data were gathered from electronic medical records. Fields collected were: demographics (age, gender, ethnicity, presence or absence of co-morbidity as defined in the Intensive Care National Audit & Research Centre (ICNARC) report on COVID-19 in critical care (Online Resource 1) and type 2 diabetes mellitus); hospital admission details (date, days of symptom onset prior to admission and radiology findings); ICU details (date of admission, mechanical ventilation during the first 24 hours, advanced respiratory support (Online Resource 1), acute physiology and chronic health evaluation (APACHE II) score and outcomes); antibiotics received and all microbiology test results to the end of the ICU admission (including any identified antimicrobial resistance).

### Definitions

Diagnostic microbiology tests were performed as per standard testing protocols within NHS laboratories at individual participating sites. Microbiology results included in the analysis were: standard culture (blood, sputum, tracheal-aspirate, bronchoalveolar lavage (BAL), urine) and validated culture-independent tests such as respiratory viral PCR (see Online Resource 2), urinary antigens and serology for *Mycoplasma pneumoniae* (IgM/ IgG). Co-infection was defined as present if a likely pathogen was identified in a clinical sample taken for diagnostic purposes. Culture results were excluded if they were considered to represent contamination or colonisation. Specifically, this applied to the following situations: blood cultures yielding common skin contaminants in a single sample (Coagulase-negative Staphylococci, *Micrococcus spp*., viridans group streptococci, *Propionibacterium spp*., *Corynebacterium spp*., *Bacillus spp*.) without a concurrent positive culture from an indwelling line tip [9-11], *Candida spp*. cultured from respiratory and urinary catheter samples [12, 13], respiratory samples yielding Gram-positive organisms typically present in the oropharyngeal flora [14], growth of *Enterococcus spp*. in a single catheter urinary specimen [15]. Radiology findings were defined based on the COVID-19 British Society of Thoracic Imaging reporting template [16]. Where both chest CT and CXR findings were available, chest CT findings were prioritised.

### Statistical analysis

Demographics, clinical and disease characteristics were described using appropriate descriptive statistics for: i) those with co-infection, and ii) those without co-infection. Characteristics of patients in the study were also compared with the patients in the Intensive Care National Audit & Research Centre (ICNARC) report on COVID-19 in critical care, 22 May 2020. The proportion of co-infection (%) was determined at three time points: on admission, within 48 hours, and during ICU admission (from day of ICU admission to ICU discharge or death in ICU). The co-infection rate was calculated per 1000 person-days based on the first co-infection detected in hospital per patient (person-time was determined from date of hospital admission to date of first co-infection, date of discharge from ICU or date of death in ICU, whichever came first for each patient). Univariate logistic regression analyses were conducted to determine the association between selected variables (age, gender, study site, ethnicity and co-morbidities) and the odds of a) developing co-infection during admission, and b) co-infection and mortality in ICU. Competing-risks regression analysis was conducted to assess if patients with co-infection had a longer length of hospital stay (from hospital admission to the end of ICU admission) than those without co-infection, with death as a competing-event. Co-pathogens were described separately for bacterial, viral and fungal infections. The proportion of bacterial co-pathogens with antimicrobial resistance was recorded.

An analysis of type of pathogens identified at different time points from admission was performed (≤48 hours and >48 hours following admission) to identify those with community vs hospital-acquired co-infection. Pathogens identified within 48 hours of hospital admission were listed by type of test performed. A sub-analysis of the hospital-acquired co-infection was performed to identify the type of pathogens detected early (3-7 days into hospital admission) and late (>7 days into hospital admission). Statistical analyses were performed using Stata MP/15.1.

## Results

Of 599 eligible patients during the study period, 254 patients with completed ICU episodes were studied (**Fig 1**).

**Fig 1:**
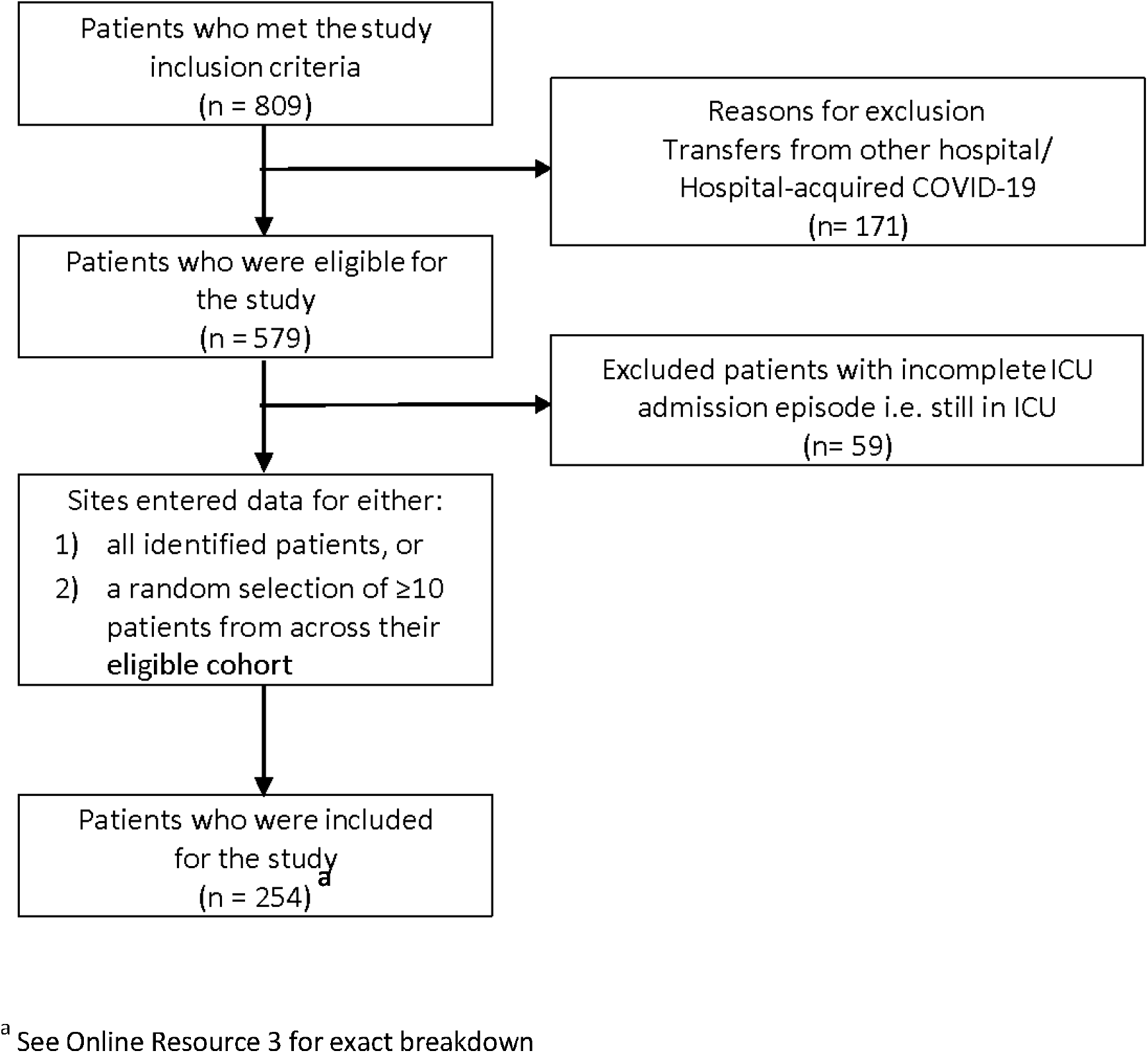
Flowchart of study population.

The median age of the study cohort was 59 years (IQR 49-69, range 19-84) and 164 (64.6%) patients were male; similar to corresponding data from the ICNARC cohort (**Table 1**)[17]. Patients were admitted to hospital between 21 Feb 2020 and 1 May 2020. The median time from onset of symptoms to admission was seven days (IQR 5-10). The median time from hospital admission to ICU admission was one day (IQR 0-2). Antibiotics were prescribed to 35 (13.8%) patients before hospital admission and to 228 (89.8%) patients within 48 hours of admission. Throughout the course of admission, 241 (94.9%) of patients received antibiotics at some point.

**Table 1:** Characteristics of study population in comparison with ICNARC data

|  | Without<br>coinfection<br>n (%) | With<br>coinfection<br>n (%) | ICNARC<br>data<sup>a</sup><br>n (%) |
| --- | --- | --- | --- |
| **Number of patients** | 171 (67.3) | 83 (32.7) | 9026 |
| **Age** |  |  | * |
| 18-49 | 47 (27.5) | 17 (20.5) |  |
| 50-64 | 51 (29.8) | 42 (50.6) |  |
| 65-74 | 47 (27.5) | 19 (22.9) |  |
| 75-84 | 26 (15.2) | 5 (6.0) |  |
| **Gender** |  |  | [N=9022] |
| Male | 106 (62.0) | 58 (69.9) | 6403 (71.0) |
| Female | 65 (38.0) | 25 (30.1) | 2619 (29.0) |
| **Ethnicity** |  |  | [N=8185] |
| White | 108 (63.2) | 44 (53.0) | 5468 (66.8) |
| Black | 13 (7.6) | 10 (12.1) | 1245 (15.2) |
| Asian | 16 (9.4) | 5 (6.0) | 797 (9.7) |
| Mixed | 3 (1.8) | 2 (2.4) | 138 (1.7) |
| Other | 4 (2.3) | 3 (3.6) | 537 (6.6) |
| <sup>§</sup>BAME | 36 (21.1) | 20 (24.1) | - |
| Unknown | 27 (15.8) | 19 (22.9) | - |
| **Co-morbidities** |  |  | [N=8777] |
| Cardiovascular | 3 (1.8) | 0 (0.0) | 42 (0.5) |
| Respiratory | 0 (0.0) | 2 (2.4) | 74 (0.8) |
| Renal | 3 (1.8) | 2 (2.4) | 144 (1.6) |
| Liver | 0 (0.0) | 0 (0.0) | 33 (0.4) |
| Metastatic disease | 0 (0.0) | 0 (0.0) | 38 (0.4) |
| Haematological malignancy | 6 (3.5) | 1 (1.2) | 144 (1.6) |
| Immunocompromised | 11 (6.5) | 2 (2.4) | 295 (3.4) |
| Type 2 diabetes mellitus | 47 (27.5) | 19 (22.9) | N/A |
| **Indicator of acute severity** |  |  |  |
| Mechanically ventilated within first 24h |  |  | 5298 (62.8)<sup>b</sup> |
| APACHE II Score, mean (SD) | 13.3 (5.6) | 14.2 (5.5) | 14.7 (5.3)<sup>c</sup> |
| PaO<sub>2</sub>/FiO<sub>2</sub> ratio (kPa), median (IQR);<br>[mmHg] | 17.2 (12.6-22.3);<br>[129 (95-168)] | 17.4 (11.8-23.7);<br>[131 (88.5-178.1)] | 15.8 (11.3-22.0)<sup>d</sup> |
| ≤ 13.3 kPa (< 100 mmHg) | 49 (28.7) | 24 (28.9) | 2982 (36.8) |
| > 13.3 and ≤ 26.7kPa (100 - 200 mmHg) | 92 (53.8) | 41 (49.4) | 3961 (48.9) |
| > 26.7 kPa (> 200 mmHg) | 30 (17.5) | 18 (21.7) | 1161 (14.3) |
| **LOS from hospital admission to the end of ICU admission (days), median (IQR)** |  |  |  |
| Survivors | 9 (4-14) | 22 (17-27) | N/A |
| Non-survivors | 7 (4-12) | 17 (11-20) |  |

<sup>a</sup> Intensive Care National Audit & Research Centre (ICNARC) report from 22 May 2020
\* Median age= 60 (51-68)
Denominators: <sup>b</sup> N=8433, <sup>c</sup> N=8648 and <sup>d</sup> N=8104

The overall median length of stay (LOS) in ICU was nine days (IQR 4-17); 10 days (IQR 4-18) for survivors and nine days (IQR 5-15) for non-survivors. One hundred and fifty-one patients (59.5%) were mechanically ventilated within 24 hours of admission, and 158 patients (62.2%) received advanced respiratory support (invasive ventilation, CPAP via trans-laryngeal tube, extracorporeal respiratory support) during admission. Of those who were discharged from ICU (n=172 patients), two patients (1.2%) died in hospital, 147 patients (85.5%) were discharged from hospital and 23 patients (13.4%) remained in hospital at the end of the study.

All patients had either a CXR (n=246 patients) and/or a chest CT scan (n= 74 patients). Classic/ probable COVID-19 radiographic changes were recorded in 209 patients (82.3%), five (2%) had normal imaging, 27 (10.6%) had indeterminate changes and 13 (5.1%) had non-COVID19 findings.

In total, co-infection was identified in 83 (32.7%) patients from hospital admission to the end of ICU stay; median time to co-infection was 9 days (IQR 6-14). The list of identified pathogens and contaminants from standard cultures (blood, BAL, sputum and tracheal aspirate) is available in Online Resource 4. On the day of admission, co-pathogens were identified in four patients (1.6%), rising to 14 (5.5%) patients within the first 48 hours of hospital admission. Fifteen pathogens were identified from 14 patients within 48 hours; 14 bacterial and one viral pathogen (**Table 2**). None of these pathogens were identified from blood culture. In a sensitivity analysis excluding the hospital which contributed a third of cases, the 48-hour co-infection rate remained similar (Online Resource 5). The commonest co-pathogen within 48 hours of hospital admission was *S. aureus*, three methicillin-susceptible (MSSA) and one methicillin-resistant *S. aureus* (MRSA) (4 patients). Two positive Mycoplasma IgG/ IgM tests in separate patients were deemed false positives and excluded from the analysis. The number of tests performed within 48 hours of hospital admission are listed in Online Resource 6, by type of tests and study site. For bacterial co-pathogens, the antimicrobial susceptibilities are described in Online Resource 7.

**Table 2:** Organisms identified within 48 hours of hospital admission

| Type of test | Pathogens | No of pathogens |
| --- | --- | --- |
| **Tracheal aspirate or sputum culture** |  |  |
|  | *Escherichia coli* | 1 |
|  | <sup>a</sup>*Pseudomonas sp* | 1 |
|  | *Pseudomonas aeruginosa* | 1 |
|  | *Enterobacter cloacae* complex (*AmpC*) | 1 |
|  | <sup>a</sup>*Staphylococcus aureus* (MSSA) | 2 |
| **BAL PCR/ culture** |  |  |
|  | <sup>b</sup>*Staphylococcus aureus* (MSSA & MRSA) | 2 |
|  | *Klebsiella pneumoniae* | 1 |
| **Other tests** |  |  |
| Pneumococcal urinary antigen test | <sup>a</sup>*Streptococcus pneumoniae* | 2 |
| MSU | *Escherichia coli* | 2 |
| Respiratory viral PCR | *Mycoplasma pneumoniae* | 1 |
|  | <sup>a</sup>*Metapneumovirus* | 1 |

<sup>a</sup> Pathogens identified on the day of admission (one out of two *Staphylococcus aureus* was from tracheal aspirate or sputum culture and one out of two pneumococcal urinary antigen tests was positive on the day of admission), total= 4
<sup>b</sup> One out of two organisms was Methicillin-resistant *Staphylococcus aureus* (MRSA). The same patient also had MRSA in pleural fluid culture after 48 hours into hospital admission.

Beyond 48 hours of hospital admission to the end of ICU stay, 124 co-pathogens were identified in 77 (30.3%) patients; 29 pathogens from Days 3 – 7 and 95 pathogens from Day 8 onwards (**Fig 2**). All were bacterial pathogens (n=122) except for two fungal organisms. The commonest co-pathogens identified were Gram-negative bacteria, including *Klebsiella spp*. (23 patients) and *Escherichia coli* (20 patients). No viral co-pathogens were detected. Of the two fungal co-pathogens, one was *Aspergillus fumigatus* from a tracheal aspirate culture obtained on Day 5 in a 54-year old male. The other was *Candida parapsilosis* from a blood culture taken at Day 7 in a 55-year old lady. Neither patient had any pre-existing co-morbidities.

**Fig 2:**
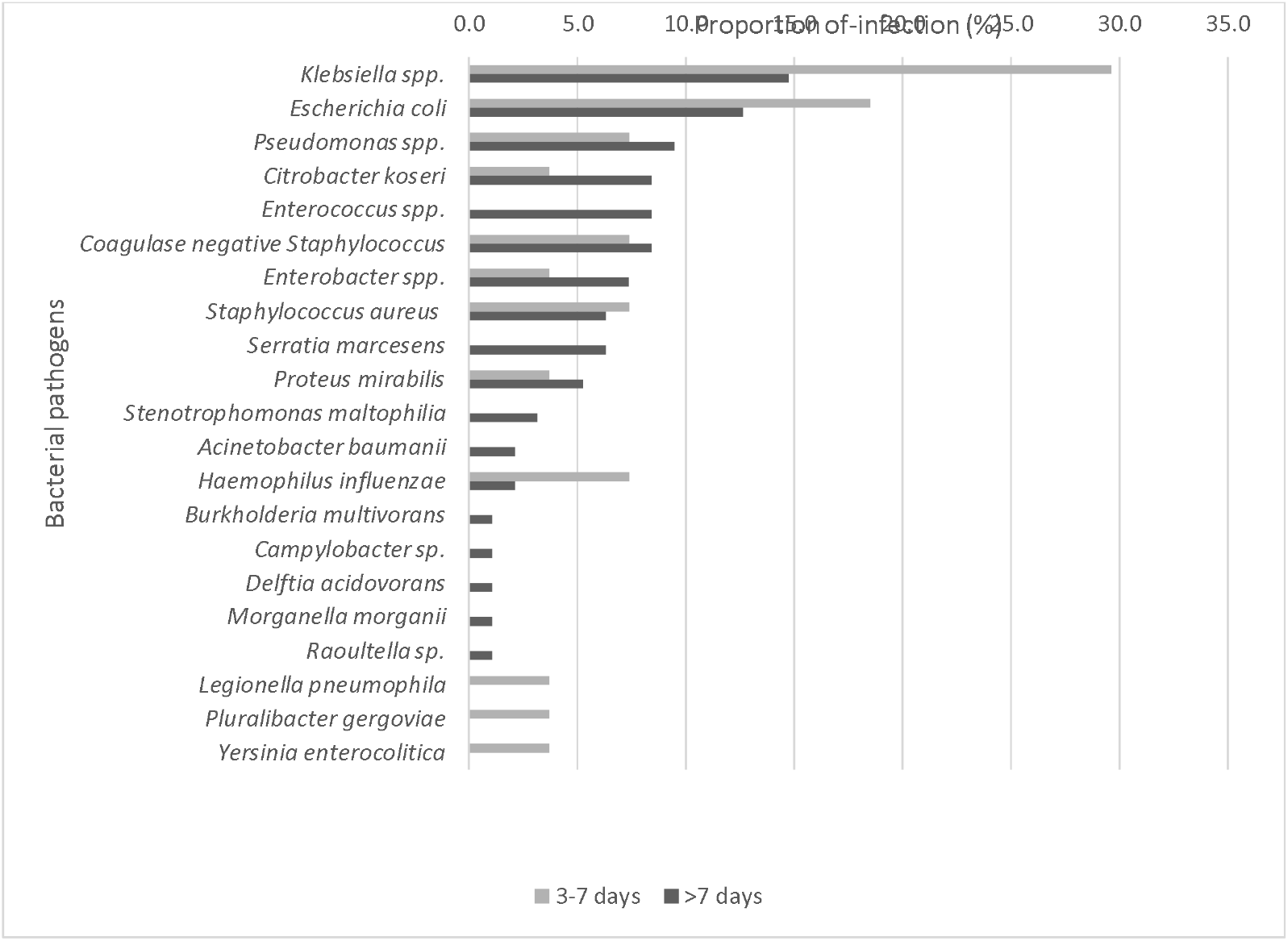
Bacterial pathogens detected after 48 hours of hospital admission; 124 pathogens detected

Reported as proportion (%) of the total number of bacterial pathogens detected within ‘3-7 days’ and ‘>7 days’ from hospital admission.

On univariate analyses, patients aged 50-64 years were more likely to have a co-infection than those aged 18-49 years. No other significant association was found (**Table 3**). Patients with co-infections were more likely to die in ICU (n=34, crude OR 1.78, 95% CI 1.03-3.08, p=0.04) and had a longer hospital LOS (measured from admission to hospital to the end of ICU admission, subhazard ratio= 0.53, 95% CI 0.39-0.71, p< 0.001) compared to those without co-infections (n=48 died).

**Table 3:** Univariate logistic regression analyses investigating the association between variables of interest and odds of developing co-infection.

|  | Crude OR<br>(95% CI) | p value |
| --- | --- | --- |
| **Number of patients** |  |  |
| **Age** |  |  |
| 18-49 | 1 (Reference) |  |
| 50-64 | 2.28 (1.14-4.53) | 0.019* |
| 65-74 | 1.12 (0.52-2.41) | 0.777 |
| 75-84 | 0.53 (0.18-1.61) | 0.263 |
| **Gender** |  |  |
| Male | 1 (Reference) |  |
| Female | 0.70 (0.40-1.23) | 0.218 |
| **Ethnicity** |  |  |
| White | 1 (Reference) |  |
| Black | 1.89 (0.77-4.62) | 0.164 |
| Asian | 0.77 (0.26-2.22) | 0.625 |
| Mixed | 1.64 (0.26-10.13) | 0.597 |
| Other | 1.84 (0.40-8.57) | 0.437 |
| <sup>a</sup>BAME | 1.36 (0.71-2.61) | 0.349 |
| Unknown | 1.73 (0.87-3.42) | 0.117 |
| **Co-morbidities** |  |  |
| Cardiovascular | - |  |
| Respiratory | - |  |
| Renal | 1.38 (0.23-8.43) | 0.725 |
| Liver | - |  |
| Metastatic disease | - |  |
| Haematological malignancy | 0.34 (0.04-2.83) | 0.316 |
| Immunocompromise | 0.36 (0.08-1.65) | 0.187 |
| Type 2 diabetes mellitus | 0.78 (0.42-1.44) | 0.434 |

\*p value of <0.05 denotes a significant difference <sup>#</sup> median and IQR
<sup>a</sup>BAME is the total of Black, Asian, Mixed and Other ethnicities

## Discussion

### Principal findings

Bacterial co-infection within 48 hours of hospital admission for COVID-19 infection in adults was uncommon; 1.6% on admission and 5.5% within 48 hours. The commonest pathogens identified within the first 48 hours of hospital admission were *Staphylococcus aureus* and *Streptococcus pneumoniae*. The proportion of pathogens detected increased with duration of ICU stay and consisted largely of Gram-negative bacteria, particularly *Klebsiella pneumoniae and Escherichia coli*. The co-infection rate >48 hours after admission was 27.0 per 1000 person-days (95% CI 21.3-34.1).

### Comparison with literature

Concern regarding co-infection during viral pandemics, specifically respiratory co-infection with a bacterial pathogen, is borne from previous experience in influenza. During the 2009 H1N1 influenza A pandemic, early co-infection rates were high; 22.5% within 72 hours of admission in adults requiring critical care [18]. In contrast, limited evidence from studies of Severe Acute Respiratory Syndrome and Middle East Respiratory Syndrome suggest lower co-infection rates (10.3 to 18.5%) [19, 20]. In COVID-19, systematic reviews based on studies predominantly from China reported low estimates (<7%) of bacterial co-infection [21-23]. In the UK, retrospective single-centre studies have observed low rates of bacterial co-infection [24-26]. Hughes et al. detected early bacterial infection (0-5 days from admission) in 3.2% of all hospitalised patients (13.5% of those requiring critical care), increasing to 6.1% throughout admission [25]. Youngs et al. reported bacterial co-infection within 48 hours of admission to ICU in 8% of patients with COVID-19 compared to 58% of patients with influenza, with no difference in the incidence of late infection between the two groups [26]. In the US, higher early bacterial co-infection rates (16.6%) were identified by Crotty et al.; respiratory cultures positive for oral bacteria flora constituted 15/25 of these cases [27]. In contrast to studies that relied on predominantly culture-based techniques, Kreitmann et al. identified early bacterial co-infection in 27.7% (13/47) of their prospective cohort of ventilated patients using a multiplex PCR assay with only one case identified by conventional culture [28]. In France, a single centre study using three multiplex PCR assays performed on respiratory specimens or nasopharyngeal swabs in addition to standard culture techniques retrospectively identified bacterial co-infection in 28% of 92 ICU admissions [29]. Variations in case definitions, diagnostic testing and geography may partly account for the differences observed between studies although overall, there is a suggestion that increased severity of disease, particularly when ICU care is required, is associated with increased rates of co-infection.

The prevalence of nosocomial infection is 20.6% and increases with duration of ICU stay [30, 31]. The rate of ventilator-acquired pneumonia is estimated at 1.2-8.5 per 1000 ventilator days and occurs in 9-27% of ventilated patients [32]. Our observed co-infection rate is relatively high, consistent with a patient cohort with long ICU stays (median 10 days) and requiring high levels of respiratory support.

Consistent with reports from other studies, the commonest co-infecting bacteria identified within 48 hours of admission was *S. aureus* [25, 28, 33]. In patients in whom early co-infection is suspected clinically, due consideration of *S. aureus* is warranted. However, the rate of *S. aureus* co-infection is markedly lower than that observed in pandemic influenza, suggesting it is a less significant issue with COVID-19 infection [18]. The predominant late pathogens observed were Gram-negative bacteria, particularly *K. pneumoniae*. These pathogens are commonly associated with hospital and ventilator-acquired pneumonia and have been reported as common co-pathogens in COVID-19 infections, particularly ICU cohorts [21, 22, 34-36]. The predominance of Gram-negative bacteria in these studies likely reflects nosocomial infection following prolonged ICU stay and empirical antibiotic use.

Viral co-pathogen was identified in one patient in our cohort; lower than the 3% (95% CI 1-6%) viral co-infection rate reported in systematic reviews and in contrast to the 20.7% viral co-detection rate reported by Kim et al. in Northern California [21, 37]. The 2019/20 influenza season in the UK ended in late March [38]. Other UK cohorts recruited during the spring wave of COVID-19 (March - May 2020) similarly reported very little or no viral co-infection [25, 36].

### Strengths and limitations

This pragmatic multicentre study provides novel data on both community-acquired and nosocomial co-infection in patients with COVID-19 requiring ICU care in England. The ICU cohort represents those with severe disease who were subject to more rigorous microbiology sampling. A key limitation of the study is its retrospective observational design subject specifically to case selection, ascertainment and sampling biases. Inclusion of consecutive eligible patients was not feasible due to pandemic workload constraints. To minimise case selection bias, participating sites submitted a random sample of their eligible cohort, although random sampling methods were not standardised. The impact of ascertainment bias due to differences in the proportion of eligible cases submitted by each institution was reduced through the participation of multiple centres. The study cohort was comparable to the ICNARC cohort except for an under-representation of patients of Black, Asian and Minority Ethnicity (BAME). Our results may not be applicable to settings with larger BAME populations. Restriction of our cohort to those with completed ICU admissions excluded: i) frailer patients in whom ICU care was deemed not appropriate, and ii) patients with very long ICU stays. Co-infection, particularly nosocomial infection, may be higher in these patients.

A second key limitation is that although results likely to represent contamination were excluded, some pathogens found in respiratory tract samples may represent colonisation rather than active co-infection. However, as sputum samples sent from ICU reflect clinical concern of lower respiratory tract infection (especially during the pandemic timeframe) and positive culture represents predominant presence of a pathogen rather than as part of mixed flora, we have taken these results to represent infection. If colonising pathogens were wrongly attributed as causing infection, the direction of bias would be towards falsely higher co-infection rates observed in our study.

Thirdly, reliance on culture dependent techniques may have falsely decreased co-infection rates. Antibiotic use prior to admission was low (13.8%), increasing the reliability of culture-based methods on admission. However, detection of pathogens later into admission would have been influenced by sampling bias and the use of empirical antibiotics. Fourthly, although seven hospitals participated in this study, one study site contributed a third of cases; observed 48-hour co-infection rate excluding this site was, however, similar to overall results.

### Implications for future work

Notwithstanding these limitations, our data indicate that early in hospitalisation, bacterial co-infection in COVID-19 is very uncommon and support the recommendations that empirical antibiotics should not be started routinely in primary care or at the point of hospital admission without clinical suspicion of bacterial infection [8]. The high rate of co-infection found late in illness among patients requiring ICU and involving nosocomial pathogens is concerning. It is plausible that reducing unnecessary early antibiotic exposure in patients with COVID-19 could reduce their risk of late, Gram negative, potentially antibiotic resistant infections [39, 40].

Since study completion, dexamethasone has been shown to decrease mortality in patients hospitalised with COVID-19 who require oxygen support or invasive mechanical ventilation [41]. Consequently, dexamethasone has become established as standard of care for these patients in many countries. This may increase the already high rate of bacterial co-infection we observed in ICU-treated patients. A high level of microbiological vigilance is recommended as part of the management of these patients. In the setting of seasonal changes in respiratory pathogens, ongoing surveillance for co-infections in patients hospitalised with COVID-19, ideally through prospective studies with standardised sampling protocols, is advised.

## Data Availability

Data are available from the corresponding author on reasonable request.

## Declarations

### Availability of data and material

Data are available from the corresponding author on reasonable request.

### Ethical approval

Ethical approval was given by HRA and REC; protocol number:20RM040, IRAS project ID:284341. Section 251 support from the Confidentiality Advisory Committee for use of anonymised NHS patient data was not required according to the temporary General Notice issued for COVID-19 purposes by the Secretary of State for Health and Social Care under the Health Service Control of Patient Information Regulations 2002.

### Authors’ contributions

All included authors fulfil the criteria of authorship; VB and HL are joint first authors for this manuscript. WSL, VB and HL had substantial contributions to the study conception and design. All authors had substantial contributions to the data acquisition. VB performed the analyses. All authors had substantial contributions to the results interpretation. VB and HL wrote the original draft. All authors revised the manuscript critically for important intellectual content, provided the final approval of the version to be published and agreed to be accountable for all aspects of the work in ensuring that questions related to the accuracy or integrity of any part of the work are appropriately investigated and resolved.

## Acknowledgement

We wish to thank Mr Glenn Hearson for building the study database on the secure online database platform (REDCAP Cloud).

## Supplementary Files

### Online Resource 1

#### Definition (based on ICNARC report on COVID-19 in critical care)^9^

**Comorbidities** must have been evident within the six months prior to critical care and documented at or prior to critical care:

- Cardiovascular: symptoms at rest
- Respiratory: shortness of breath with light activity or home ventilation
- Renal: renal replacement therapy for end-stage renal disease
- Liver: biopsy-proven cirrhosis, portal hypertension or hepatic encephalopathy
- Metastatic disease: distant metastases
- Haematological malignancy: acute or chronic leukaemia, multiple myeloma or lymphoma
- Immunocompromise: chemotherapy, radiotherapy or daily high dose steroid treatment in previous six months, HIV/AIDS or congenital immune deficiency
- Type II diabetes mellitus

**Mechanical ventilation during the first 24 hours** was identified by the recording of a ventilated respiratory rate, indicating that all or some of the breaths or a portion of the breaths (pressure support) were delivered by a mechanical device. This usually indicates invasive ventilation; BPAP (bi-level positive airway pressure) would meet this definition but CPAP (continuous positive airway pressure) does not.

**Advanced respiratory support** was defined as invasive ventilation, BPAP via trans-laryngeal tube or tracheostomy, CPAP via trans-laryngeal tube, extracorporeal respiratory support.

### Online Resource 2

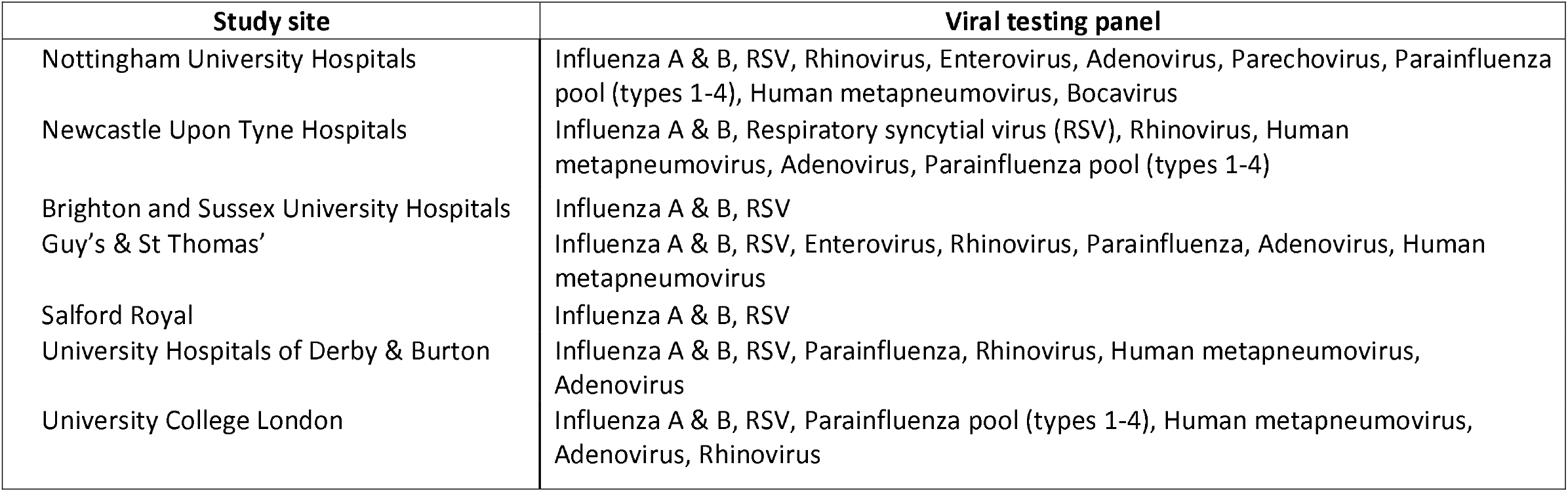

### Online Resource 3

| Study site | Met inclusion criteria<sup>a</sup> | Still in ICU<sup>b</sup> | Transfers from other hospital/ Hospital-acquired COVID-19<sup>c</sup> | Eligible<sup>d</sup> | Entered into database (% of those eligible) |
| --- | --- | --- | --- | --- | --- |
| Nottingham University Hospitals | 97 | 11 | 1 | 85 | 79 (92.9) |
| Newcastle Upon Tyne Hospitals | 100 | 4 | 20 | 76 | 48 (63.2) |
| Brighton and Sussex University Hospitals | 58 | 8 | 4 | 46 | 45 (64.3) |
| Guy's & St Thomas' NHS Foundation Trust | 316 | 0 | 103 | 213 | 34 (16.0) |
| Salford Royal | 46 | 3 | 11 | 32 | 22 (68.8) |
| University Hospitals of Derby & Burton | 54 | 1 | 0 | 53 | 16 (30.2) |
| University College Hospitals London | 138 | 32 | 32 | 74 | 10 (13.5) |
| **Total** | **809** | **59** | **171** | **579** | **254 (43.9)** |

d= a- (b+c)

### Online Resource 4

**Table S1:**
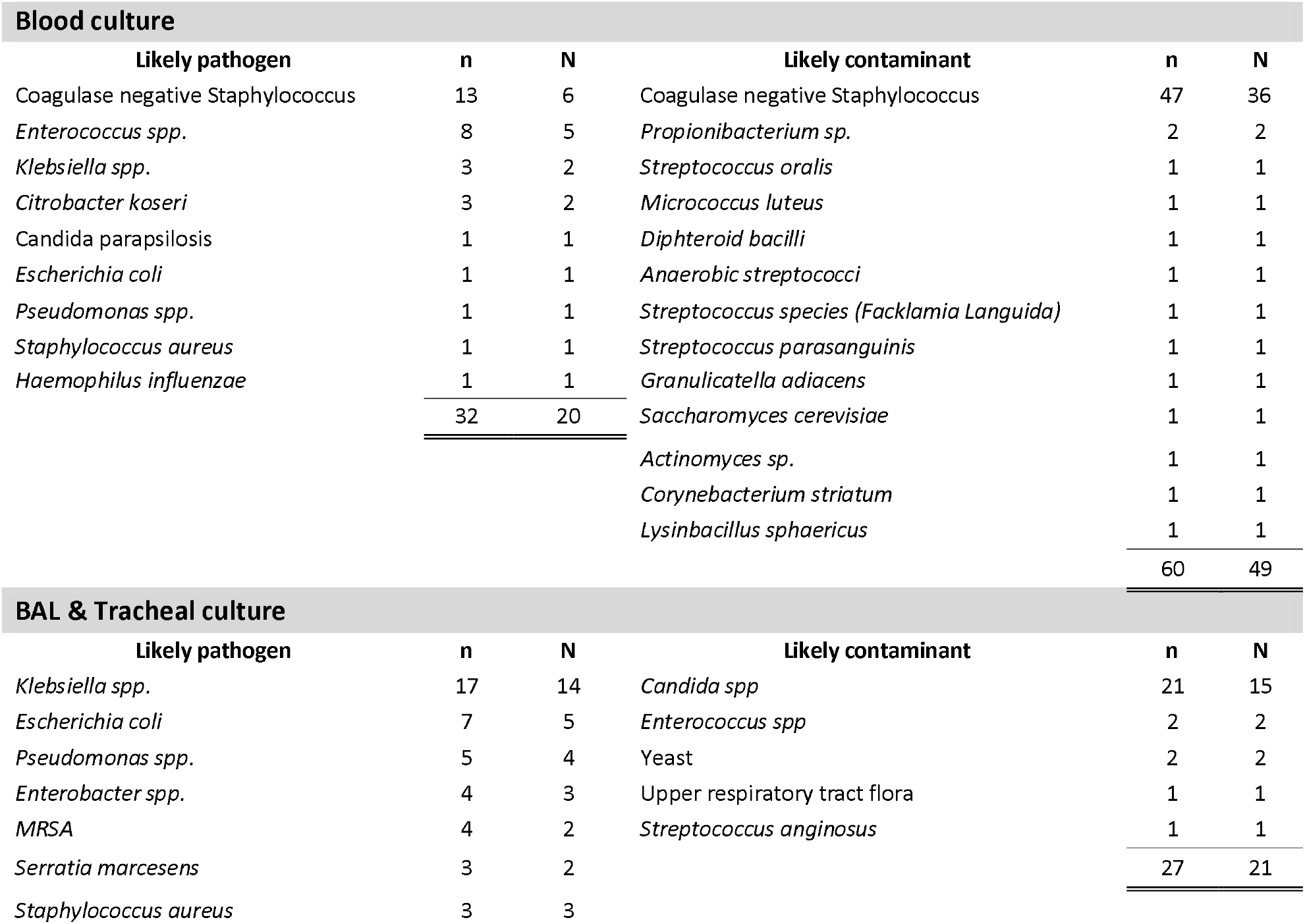
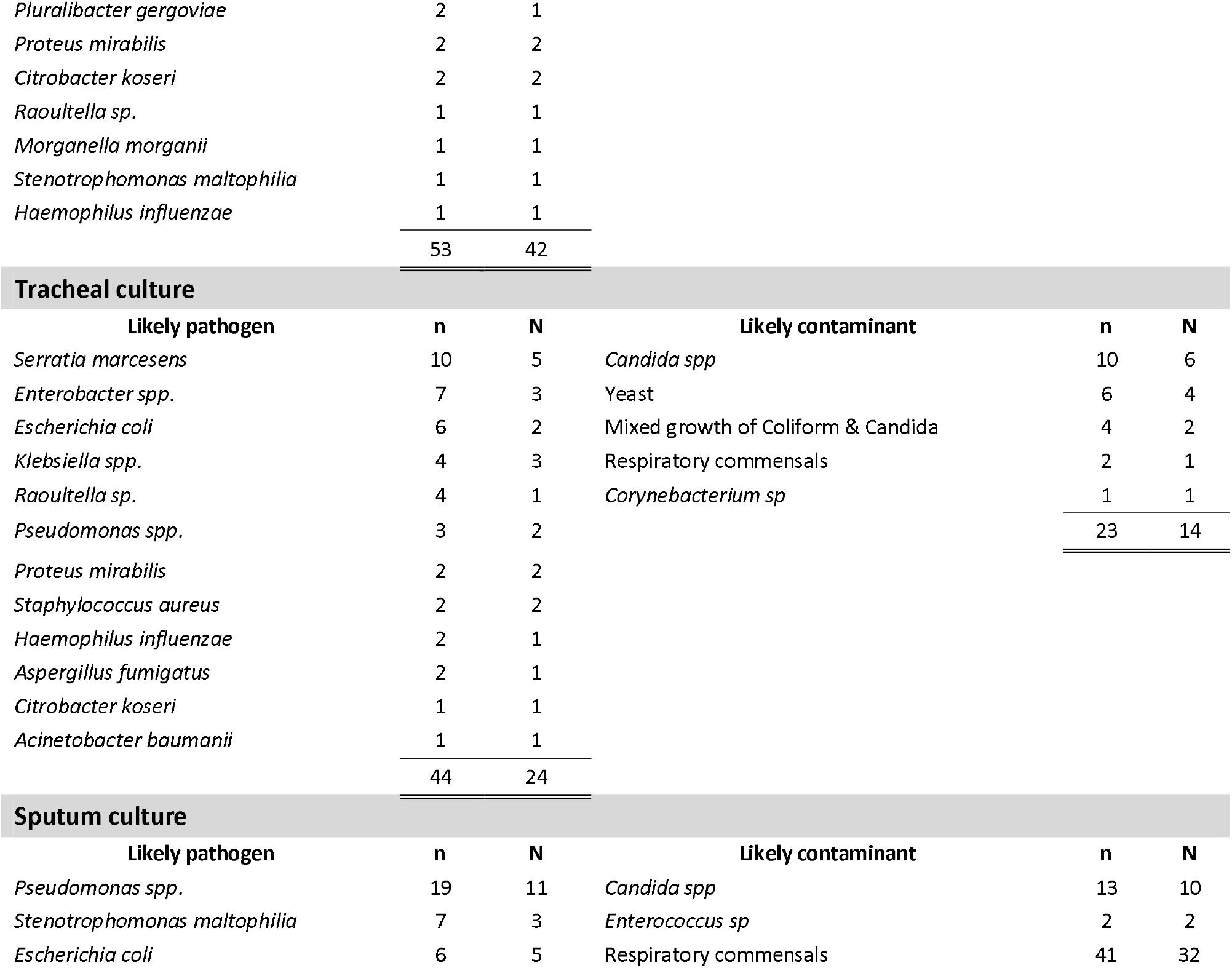
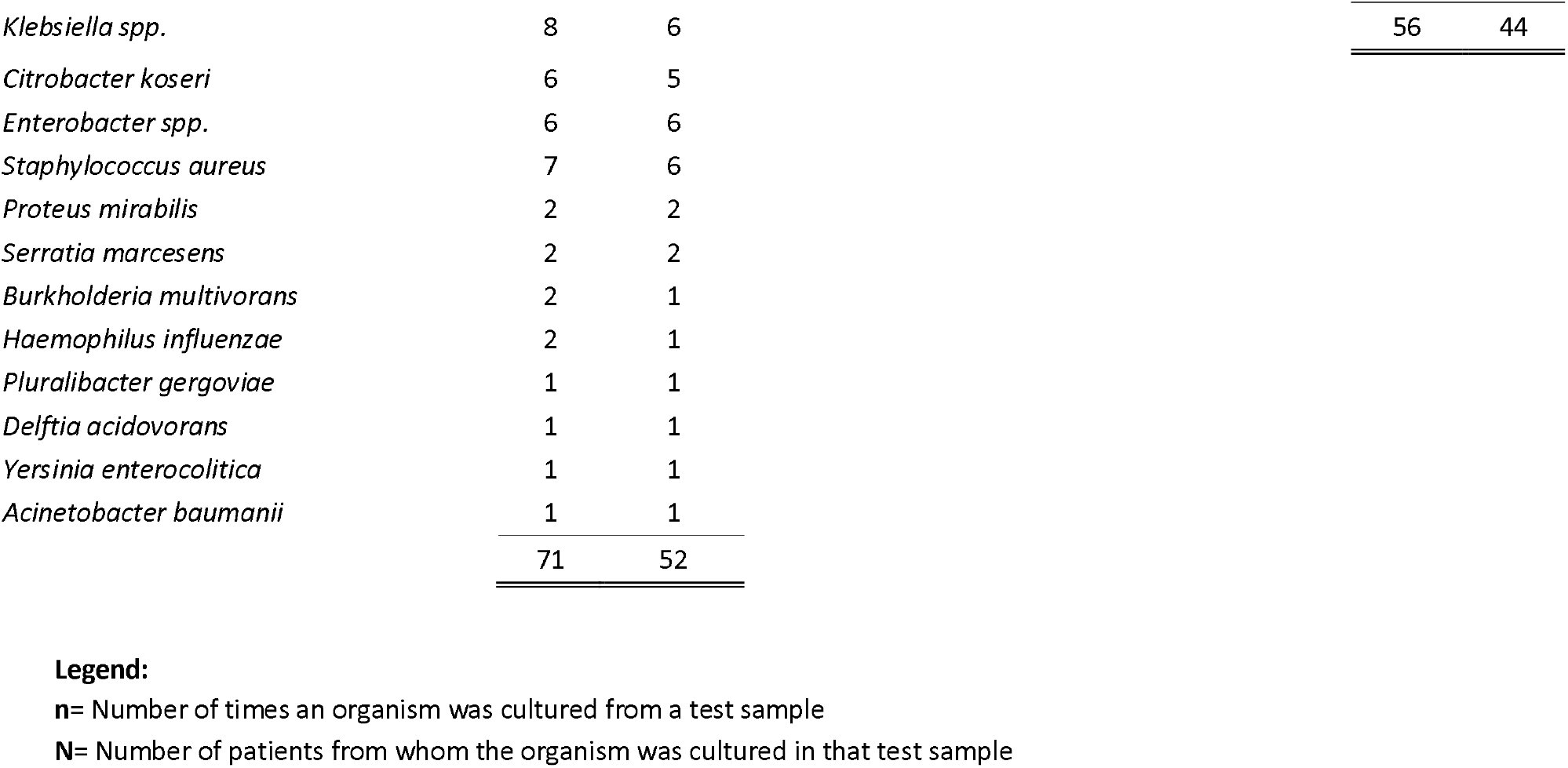
Results and classification as likely pathogen or contaminant among positive cultures taken from patients

### Online Resource 5

|  | Co-infection rate within 48 hours<br>(1000 person-days) (95% CI) |
| --- | --- |
| Overall | 28.2 (16.7-47.7) |
| Excluding Nottingham University Hospitals | 32.0 (17.7-57.8) |

### Online Resource 6

| Type of tests | On admission |  | After admission<br>(within 48 hours) |  | Overall |  |
| --- | --- | --- | --- | --- | --- | --- |
|  | n | N | n | N | n | N |
| Blood culture | 223 | 174 | 89 | 46 | 312 | 220 |
| BAL PCR/ culture, sputum culture,<br>tracheal culture | 18 | 15 | 34 | 31 | 52 | 46 |
| Urinary pneumococcal antigen | 25 | 25 | 55 | 51 | 80 | 76 |
| Urinary legionella antigen | 36 | 34 | 55 | 54 | 91 | 88 |
| Respiratory viral PCR | 119 | 106 | 32 | 22 | 151 | 128 |

**n**= Number of tests done
**N**= Number of patients who had the test

### Online Resource 7

|  | AMR | No AMR | Unknown | Total | Resistance n (%) |
| --- | --- | --- | --- | --- | --- |
| *Klebsiella spp.* | 18 | 3 | 2 | 23 | Amoxicillin, 16 (88.9); Co-amoxiclav, 9 (50); Cefuroxime, 7 (38.9); Piperacillin/Taz, 5 (27.8); Meropenem, 1 (5.6) |
| *Escherichia coli* | 11 | 2 | 0 | 20 | Amoxicillin, 8 (72.7); Co-amoxiclav, 5 (45.5); Meropenem, 1 (9.1); Ertapenem, 1 (9.1) |
| *Enterobacter spp.* | 7 | 2 | 0 | 9 | Amoxicillin, 6 (85.7); Co-amoxiclav, 6 (85.7); Cefuroxime, 2 (28.6); Cefadroxil, 1 (14.3); Ceftazidime, 1 (14.3); Meropenem, 1 (14.3); Gentamicin, 1 (14.3) |
| *Pseudomonas spp.* | 7 | 4 | 2 | 13 | Ciprofloxacin, 2 (28.6); Ceftazidime, 2 (28.6); Piperacillin/Taz, 5 (71.4); Meropenem, 3 (42.9); Gentamicin, 1 (14.3); Amikacin, 1 (14.3); Ticarcillin/ clavulanate, 1 (14.3) |
| *Serratia marcescens* | 7 | 0 | 0 | 7 | Amoxicillin, 1 (100); Co-amoxiclav, 6 (85.7); Cefuroxime, 1 (14.3); Piperacillin/Taz, 1 (14.3) |
| *Citrobacter koseri* | 5 | 3 | 1 | 9 | Amoxicillin, 5 (100); Co-amoxiclav, 1 (20); Piperacillin/Taz, 1 (20); Meropenem, 1 (20) |
| *Staphylococcus aureus* | 4 | 7 | 0 | 11 | Flucloxacillin, 1 (25); Doxycycline, 2 (50); Clarithromycin, 3 (75); Clindamycin, 1 (25) |
| *Haemophilus influenzae* | 3 | 0 | 1 | 4 | Amoxicillin, 3 (100); Co-amoxiclav, 3 (100); Cefuroxime, 2 (66.7); Doxycycline, 1 (33.3) |
| *Acinetobacter baumannii* | 1 | 1 | 0 | 2 | Ceftazidime, 1 (100) |
| *Burkholderia multivorans* | 1 | 0 | 0 | 1 | Gentamicin, 1 (100); Meropenem, 1 (100); Ceftolozane/Tazobactam, 1 (100) |
| *Enterococcus spp.* | 1 | 2 | 5 | 8 | Amoxicillin, 1 (100); Gentamicin, 1 (100) |
| *Morganella morganii* | 1 | 0 | 0 | 1 | Amoxicillin, 1 (100); Co-amoxiclav, 1 (100); Cefuroxime, 1 (100); Piperacillin/Taz, 1 (100) |
| *Raoultella sp.* | 1 | 0 | 0 | 1 | Amoxicillin, 1 (100); Piperacillin/Taz, 1 (100) |
| *Yersinia enterocolitica* | 1 | 0 | 0 | 1 | Amoxicillin, 1 (100); Co-amoxiclav, 1 (100) |
| **Total** | **68** | **24** | **11** | **110** |  |

